# Evaluating AI Proficiency in Nuclear Cardiology: Large Language Models take on the Board Preparation Exam

**DOI:** 10.1101/2024.07.16.24310297

**Authors:** Valerie Builoff, Aakash Shanbhag, Robert JH Miller, Damini Dey, Joanna X. Liang, Kathleen Flood, Jamieson M. Bourque, Panithaya Chareonthaitawee, Lawrence M. Phillips, Piotr J Slomka

## Abstract

**Background:** Previous studies evaluated the ability of large language models (LLMs) in medical disciplines; however, few have focused on image analysis, and none specifically on cardiovascular imaging or nuclear cardiology.

**Objectives:** This study assesses four LLMs - GPT-4, GPT-4 Turbo, GPT-4omni (GPT-4o) (Open AI), and Gemini (Google Inc.) - in responding to questions from the 2023 American Society of Nuclear Cardiology Board Preparation Exam, reflecting the scope of the Certification Board of Nuclear Cardiology (CBNC) examination.

**Methods:** We used 168 questions: 141 text-only and 27 image-based, categorized into four sections mirroring the CBNC exam. Each LLM was presented with the same standardized prompt and applied to each section 30 times to account for stochasticity. Performance over six weeks was assessed for all models except GPT-4o. McNemar’s test compared correct response proportions.

**Results:** GPT-4, Gemini, GPT4-Turbo, and GPT-4o correctly answered median percentiles of 56.8% (95% confidence interval 55.4% - 58.0%), 40.5% (39.9% - 42.9%), 60.7% (59.9% - 61.3%) and 63.1% (62.5 – 64.3%) of questions, respectively. GPT4o significantly outperformed other models (p=0.007 vs. GPT-4Turbo, p<0.001 vs. GPT-4 and Gemini). GPT-4o excelled on text-only questions compared to GPT-4, Gemini, and GPT-4 Turbo (p<0.001, p<0.001, and p=0.001), while Gemini performed worse on image-based questions (p<0.001 for all).

**Conclusion:** GPT-4o demonstrated superior performance among the four LLMs, achieving scores likely within or just outside the range required to pass a test akin to the CBNC examination. Although improvements in medical image interpretation are needed, GPT-4o shows potential to support physicians in answering text-based clinical questions.

## INTRODUCTION

In recent years, rapid advancements in large language models (LLMs) have prompted investigations into their potential applications across various fields, including medicine^1,2^. To explore the potential utility of LLMs in medical contexts, studies have evaluated the performance of these models on the United States Medical Licensing Exam (USMLE)^3^ and medical subspecialty board examinations^4–9^. Successfully answering these exam questions suggests that LLMs may reach the baseline knowledge levels expected of physicians, indicating their potential as educational aids that can help elucidate complex concepts and provide explanations for correct answers^3^. However, the application of LLMs in supporting diagnostic processes and medical decision-making requires cautious interpretation and further validation. Particularly, the capacity of LLMs to analyze and interpret multimodal questions that include medical images—a critical component in cardiovascular imaging— remains underexplored. While some studies have begun to evaluate image analysis capabilities in individual LLMs, comprehensive comparisons across different models and over time are lacking ^10–14^. This research gap underscores the need for more in-depth investigation into the image interpretation abilities of different LLMs, especially for diagnostics that integrate multimodal data, such as nuclear cardiology.

Nuclear cardiology presents a unique challenge for LLMs due to its intricate diagnostic processes and reliance on nuanced interpretations of both imaging and textual data. Unlike general medical exams such as the USMLE, the specialized knowledge for the nuclear cardiology exam is derived from a narrower selection of medical journals. This could pose challenges for models trained using reinforcement learning techniques. In the United States, physicians are required to pass the Certification Board of Nuclear Cardiology (CBNC) exam to obtain and maintain certification in this field. The American Society of Nuclear Cardiology (ASNC) Board Preparation Examinations, which align with CBNC standards^15^, are frequently used for exam preparation. Consequently, achieving a certain percentage of correct responses on an ASNC board preparation exam may serve as a proxy for the minimum knowledge level deemed acceptable for passing the CNBC examination. However, there is no established passing score for these preparation questions, and the precise threshold correlating to a passing score on the CBNC exam remains undetermined.

The goal of this study was to evaluate the proficiency of four state-of-the-art LLM chatbots—GPT-4, GPT-4 Turbo, GPT-4omni (GPT-4o) (Open AI) and Gemini (Google Inc.) — in answering both multimodal (text and image-based) and text-only (non-image-based) questions from the 2023 ASNC Board Preparation Examination. We separated exam questions into four key sections that mirrored the CBNC certification exam’s structure. We tested the models under standardized conditions, repeating the tests 30 times for all four LLMs. Additionally, GPT-4, Gemini and GPT-4 Turbo were tested progressively over a 6-week time interval with a single manual attempt. This approach enabled the identification of performance variability between consecutive attempts and the observation of potential performance improvement or degradation over time, revealing the strengths and weaknesses of each LLM. Ultimately, by evaluating the proficiency of LLMs, our study evaluated their potential utility as educational tools or assistive technologies within the medical community.

## METHODS

### Questions dataset

The study utilized multiple-choice questions from the 2023 ASNC Board Preparation Exam. The questions were developed by expert Nuclear Cardiology faculty for ASNC’s Board Exam Preparation Course participants preparing for the CBNC exam. None of the board preparation exam questions are used in the actual CBNC exam; however, the questions were constructed based on the CBNC exam content outline^15^. ASNC granted permission to use these questions for this study and provided the correct answers. The 2023 ASNC Board Preparation Exam comprises 168 questions, all of which were used for this study. Ethical/IRB approval was not obtained because there were no subjects involved in this study.

### Question classification

In the study, questions were categorized by topic into four sections according to ASNC guidelines: “1-Physics, Instrumentation, Radionuclides, and Radiation Safety”, “2-Acquisition and Quality Control, Gated SPECT, Artifact Recognition, and MUGA”, “3-Test Selection, Stress and Nuclear Protocols Interpretation, Appropriate Use, and Risk Stratification”, and “4-Cardiac PET, Multimodality Imaging, Cardiac Amyloidosis, Cases with the Experts: PET and SPECT.” These sections align with the structure of the CBNC certification exam, which includes “Selection of Nuclear Cardiology Imaging Tests”, “Performance of Nuclear Cardiology Imaging Tests (including instrumentation, protocols and processing)”, “Interpretation of nuclear cardiology imaging tests”, and “Radiation safety and management of radiopharmaceuticals.”^15^

Questions were also categorized by the presence or absence of images in the questions or answer choices. Overall, the dataset comprised 27 image-based questions, and 141 text-only questions, that did not include or require image interpretation. The distribution of questions across the sections is displayed in the **Graphical Abstract**.

### Model performance and data collection

GPT-4^16,17^, −4 Turbo and -Omni are Transformer-style models^18^ pre-trained to predict the next token in a document, using both publicly available data (such as internet data) and data licensed from third-party providers to OpenAI, San Francisco, CA, USA. The models are then fine-tuned using Reinforcement Learning from Human Feedback^19^. Given both the competitive landscape and the safety implications of large-scale models like GPT-4, there are no official details about the architecture (including model size), hardware, training compute, dataset construction, training method, or similar but are estimated to be in the order based on eight models with 220 billion parameters each, for a total of about 1.76 trillion^17^ parameters connected through a mixture model. Gemini (Google, Mountain View, CA, USA) is also trained on publicly available and licensed data, and has no disclosure of training compute or learnable parameters discussed and are estimated to be 137 billion^20^. The GPT-4 model was trained on data available up to September 2021. The GPT-4 Turbo with Vision Preview model received training on data available up to April 2023. Google’s Gemini 1.0 Pro with Vision was trained on data available through February 2023. GPT-4o was trained on data available through October 2023.

Responses from GPT-4, Gemini, GPT-4 Turbo, and GPT-4o were collected in two stages. In the first stage, singular test attempt responses from GPT-4, Google Gemini, and GPT-4 Turbo were collected six weeks apart in April 2024 and June 2024 to evaluate time-progressive model performance. GPT-4o was excluded from this stage of testing as it was released on May 13, 2024, after the first stage had commenced. In the second stage, to account for model stochasticity and to assess models’ performance variability between attempts, each model was applied to the examination 30 times between May 22, 2024 and May 30, 2024, in accordance with the availability of token resources and application programming interface (API) request limits.

Azure OpenAI studio within the Azure cloud ecosystem^21^ was utilized to automatically run API requests with dedicated provisioned deployments within West US for testing with 30 repetitions for all GPT versions (GPT-4, GPT-4 Turbo, and GPT-4o). The analysis used GPT default parameters controlling randomness for repetitive answers or creativeness and selection of likelier tokens specifically, utilizing a temperature of 0.7 and top_p of 0.95. All Google Gemini exam attempts, as well as singular exam attempts made in the time-progressive testing stage of the study, were repeated manually through the chat playground due to the API client restrictions for Google AI studio and paid service token restrictions in the region of use. During manual testing of each LLM, text-only questions were pasted into the respective text input fields, and image-based questions were uploaded individually as PNG files.

All questions were presented sequentially as they appear on the exam. Each LLM was presented the same standardized prompt:

> *“Please answer the following questions. Provide me only with the letters that correspond to each first most likely and second most likely answers. The first most likely and second most likely answers cannot be the same. Export only the first most likely and second most likely alphabetical answers and their corresponding question numbers as an excel sheet.”*

To reduce bias from the models potentially recalling previous inputs, the “history and model training” setting was turned off. Additionally, the session was refreshed, or a new session was started whenever the maximum token limit was reached.

### Exam Scoring

All questions had 1 correct answer. The alphabetical multiple-choice option (A, B, C, D, etc.) determined by the model was compared to the corresponding correct answer provided by ASNC. All questions were weighted equally when calculating the total score.

### Statistical Analyses

Categorical variables are presented as count and relative frequencies (percentages). To account for model variability between attempts, the test was administered 30 times to each model across all four sections of the exam. Median and 95% confidence intervals (CIs) over the resulting percentiles are reported. McNemar’s test was used to evaluate differences in the proportion of correct responses between LLMs and to compare the change in performance of a single LLM over time. A two-tailed p-value of <0.05 was considered statistically significant. All statistical analyses were performed with Python 3.11.5 (Python Software Foundation, Wilmington, DE, USA).

## RESULTS

### Overall performance

Figure 1 shows the overall performance of the models on all 168 questions of the 2023 ASNC Board Preparation Exam. GPT-4, Gemini, GPT4-Turbo, and GPT-4o correctly answered median percentiles of 56.8% (95% CI 55.4% - 58.0%), 40.5% (39.9% - 42.9%), 60.7% (59.9% - 61.3%) and 63.1% (62.5% – 64.3%) of the questions, respectively. Overall, GPT-4o significantly outperformed the other models (p=0.007 vs. GPT-4 Turbo and p<0.001 vs. GPT-4 and Gemini). There were significant differences between GPT-4, Gemini, and GPT-4 Turbo on overall performance (p<0.001 for all).

**Figure 1.**
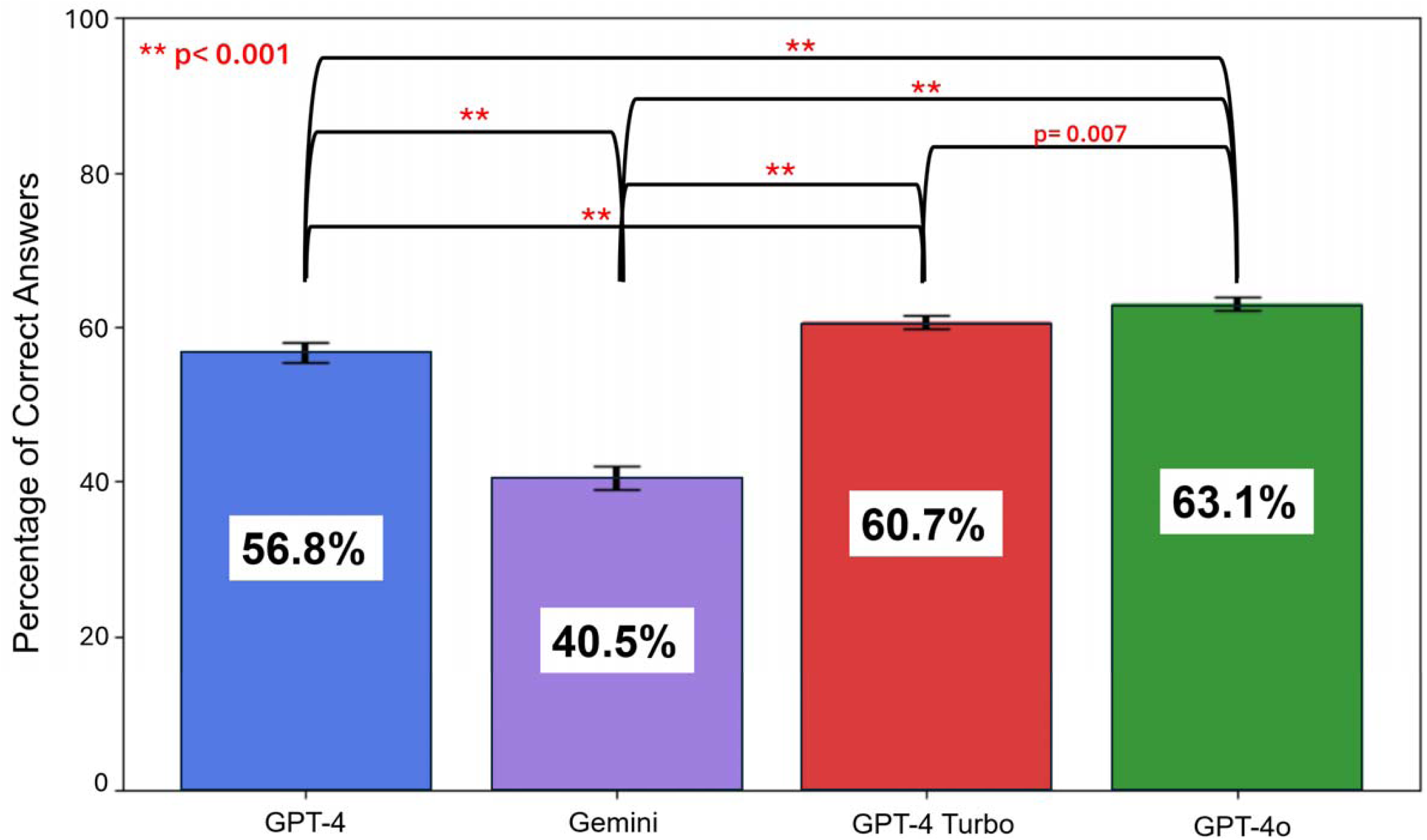
Overall Performance of LLMs. Models were applied 30 times to all 168 questions of the 2023 American Society of Nuclear Cardiology (ASNC) Board Preparation Exam. Results are reported as median percentiles of the 30 attempts, with their 95% confidence intervals. LLM – large language model

### Comparison by section type

A sectional analysis of the models’ performance is displayed in **Table 1** as the median percentage of correct answers across 30 test attempts. In Section 1 (Physics, Instrumentation, Radionuclides, and Radiation Safety), GPT-4, Gemini, GPT-4 Turbo and GPT-4o correctly answered 63.0% (58.0% - 64.0%), 38.0% (36.0% - 40.0%), 62.0% (61.2% - 62.8%) and 73.0% (71.2% - 74.9%) of questions, respectively. The proportions of correct responses were significantly different among the four models (p<0.001 for all, except for p=0.024 for GPT-4 vs. GPT-4 Turbo).

**Table 1:** Performance per section.

| Test | Section 1:<br>Physics,<br>Instrumentation,<br>Radionuclides,<br>and Radiation<br>Safety | p-value | Section 2:<br>Acquisition and<br>Quality Control,<br>Gated SPECT,<br>Artifact<br>Recognition, and<br>MUGA | p-value | Section 3:<br>Test Selection,<br>Stress and<br>Nuclear Protocols<br>Interpretation,<br>Appropriate Use,<br>and Risk<br>Stratification | p-value | Section 4:<br>Cardiac PET,<br>Multimodality<br>Imaging,<br>Cardiac<br>Amyloidosis,<br>Cases with the<br>Experts: PET<br>and SPECT | p-value |
| --- | --- | --- | --- | --- | --- | --- | --- | --- |
| GPT-4<br>(4) | 63.0%<br>(58.0% - 64.0%) | p< 0.001<br>(4 vs. G) | 44.4%<br>(41.1% - 46.7%) | p= 0.877<br>(4 vs. G) | 67.5%<br>(66.0% - 69.0%) | p< 0.001<br>(4 vs. G) | 58.0%<br>(56.7% - 59.3%) | p< 0.001<br>(4 vs. G) |
| Gemini<br>(G) | 38.0%<br>(36.0% - 40.0%) | p= 0.024<br>(4 vs. 4T) | 42.2%<br>(41.3% - 43.2%) | p< 0.001<br>(4 vs. 4T) | 40.0%<br>(37.5% - 42.5%) | p= 0.044<br>(4 vs. 4T) | 45.5%<br>(44.3% - 46.6%) | p= 0.222<br>(4 vs. 4T) |
| GPT-4 Turbo<br>(4T) | 62.0%<br>(61.2% - 62.8%) | p< 0.001<br>(G vs. 4T) | 53.3%<br>(51.1% - 55.6%) | p< 0.001<br>(G vs. 4T) | 68.8%<br>(67.5% - 70.0%) | p< 0.001<br>(G vs. 4T) | 57.6%<br>(56.1% - 60.6%) | p< 0.001<br>(G vs. 4T) |
| GPT-4o<br>(4o) | 73.0%<br>(71.2% - 74.9%) | p< 0.001<br>(4o vs. 4T) | 55.6%<br>(54.2% - 56.9%) | p= 0.306<br>(4o vs. 4T) | 60.0%<br>(57.4% - 62.5%) | p< 0.001<br>(4o vs. 4T) | 63.6%<br>(62.2% - 65.0%) | p< 0.001<br>(4o vs. 4T) |
|  |  | p< 0.001<br>(4o vs. G) |  | p< 0.001<br>(4o vs. G) |  | p< 0.001<br>(4o vs. G) |  | p< 0.001<br>(4o vs. G) |
|  |  | p< 0.001<br>(4o vs. 4) |  | p< 0.001<br>(4o vs. 4) |  | p< 0.001<br>(4o vs. 4) |  | p< 0.001<br>(4o vs. 4) |

\* Values are presented as median percentiles and 95% confidence intervals of 30 attempts at the exam. SPECT - single photon emission computed tomography ; PET – positron emission tomography; MUGA – multiple-gated acquisition

Performance of GPT-4 Turbo (53.3% [51.1%-55.6%]) and GPT-4o (55.6% [54.2% - 56.9%]) were both superior to GPT-4 (44.4% [41.1%-46.7%]) and Gemini (42.2% [41.3%-43.2%]) in Section 2 (Acquisition and Quality Control, Gated SPECT, Artifact Recognition, and MUGA), with p<0.001 for all comparisons. In this section, similar performance was observed between GPT-4 and Gemini (p=0.877) as well as between GPT-4 Turbo and GPT-4o (p=0.306).

In Section 3 (Test Selection, Stress and Nuclear Protocols Interpretation, Appropriate Use, and Risk Stratification), GPT-4, Gemini, GPT4-Turbo and GPT-4o correctly answered 67.5% (66.0% - 69.0%), 40.0% (37.5% - 42.5%), 68.8% (67.5% - 70.0%) and 60.0% (57.4% - 62.5%) of questions, respectively. The proportion of correct responses was significantly different among the four models (p<0.001 for all, except for p=0.044 for GPT-4 vs. GPT-4 Turbo).

In Section 4 (Cardiac PET, Multimodality Imaging, Cardiac Amyloidosis, Cases with the Experts: PET and SPECT), GPT-4o correctly answered 63.6% (62.2% - 65.0%) of questions, significantly outperforming the other models (p<0.001 for all). GPT-4 Turbo (57.6% [56.1% - 60.6%]) and GPT-4 (58.0% [56.7% - 59.3%]) significantly outperformed Gemini (45.5% [44.3% - 46.6%], p<0.001 and p<0.001, respectively). GPT-4 Turbo and GPT-4 demonstrated similar performance on Section 4 of the exam (p=0.222). Gemini was unable to answer two image-based questions from Section 4, which require subjective clinical evaluations, because it is not trained to answer questions that request medical advice^22,23^ (**Supplemental Figure 1 and 2**). Thus, these questions were marked as incorrect for Gemini.

**Figure 2.**
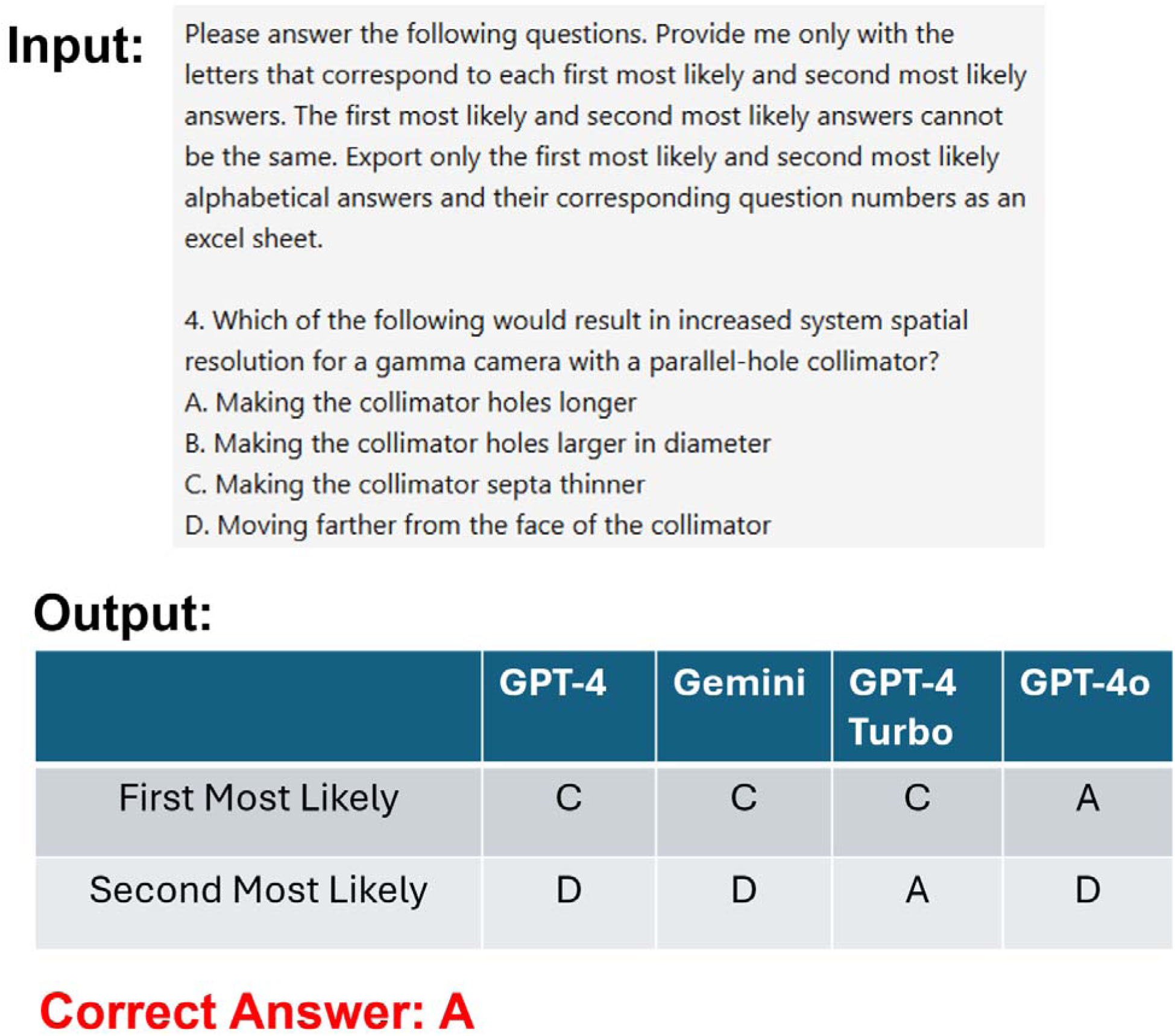
Section 1 Example Problem. The input and the three models’ output for a text-only question from Section 1 (Physics, Instrumentation, Radionuclides, and Radiation Safety) of the 2023 ASNC Board Preparation Exam. A first most likely and second most likely answer was provided by the models. Only the first most likely answer was scored and compared to the correct answer. GPT-4o was able to answer correctly, whereas GPT-4, Gemini and GPT-4 Turbo provided incorrect answers for 29 of 30 attempts. ASNC – American Society of Nuclear Cardiology

### Comparison of text-only questions

When considering text-only questions that did not include or require image interpretation, across all sections, GPT-4o (66.7% [65.7% - 67.7%]) significantly outperformed GPT-4 (59.2% [58.2% - 60.6%], p<0.001), Gemini (44.7% [44.0% - 46.1%], p<0.001) and GPT-4 Turbo (62.4% [62.1% - 63.8%], p = 0.001) (**Table 2**). Figure 2 displays a text-only example question from Section 1 of the exam, with an outcome aligning with the overall text-only question results. In this scenario, GPT-4o answered correctly, whereas GPT-4, Gemini and GPT-4 Turbo provided incorrect answers for 29 out of 30 attempts.

**Table 2:** Performance on image and no image questions.

| Test | Image Questions | p-value | No Image Questions | p-value |
| --- | --- | --- | --- | --- |
| GPT-4 (4) | 40.7%<br>(37.0% - 44.4%) | p< 0.001<br>(4 vs. G) | 59.2%<br>(58.2% - 60.6%) | p< 0.001<br>(4 vs. G) |
| Gemini (G) | 22.2%<br>(20.4% - 25.9%) | p= 0.885<br>(4 vs. 4T) | 44.7%<br>(44.0% - 46.1%) | p< 0.001<br>(4 vs. 4T) |
| GPT-4 Turbo (4T) | 44.4%<br>(40.7% - 48.1%) | p< 0.001<br>(G vs. 4T) | 62.4%<br>(62.1% - 63.8%) | p< 0.001<br>(G vs. 4T) |
| GPT-4o (4o) | 44.4%<br>(42.2% - 46.7%) | p= 0.821<br>(4o vs. 4T)<br>p< 0.001<br>(4o vs. G)<br>p= 0.956<br>(4o vs. 4) | 66.7%<br>(65.7% - 67.7%) | p= 0.001<br>(4o vs. 4T)<br>p< 0.001<br>(4o vs. G)<br>p< 0.001<br>(4o vs. 4) |

\*Values are presented as median percentiles and 95% confidence intervals of 30 attempts at the exam.

Additionally, there were significant differences between GPT-4, Gemini and GPT-4 Turbo in this category (p<0.001 for all) **(Table 2)**.

### Comparison of image-based questions

When considering questions that contained images in the questions or answers, GPT-4, Gemini, GPT-4 Turbo and GPT-4o correctly answered 40.7% (37.0% - 44.4%), 22.2% (20.4% - 25.9%), 44.4% (40.7% - 48.1%) and 44.4% (42.2% - 46.7%) questions, respectively (**Table 2**). The proportion of correct responses was similar among GPT-4 and GPT-4o (p= 0.956), GPT-4 and GPT-4 Turbo (p=0.885) and GPT-4o and GPT-4 Turbo (p=0.821). Gemini exhibited significantly worse performance compared to the other three models (p<0.001 for all). Figure 3 displays a text-only example question from Section 3 of the exam, with an outcome aligning with the overall image-based question results. In this scenario, GPT-4, GPT-4 Turbo and GPT-4o answered correctly in 17 out of 30 attempts, whereas Gemini provided incorrect answers in all attempts.

**Figure 3.**
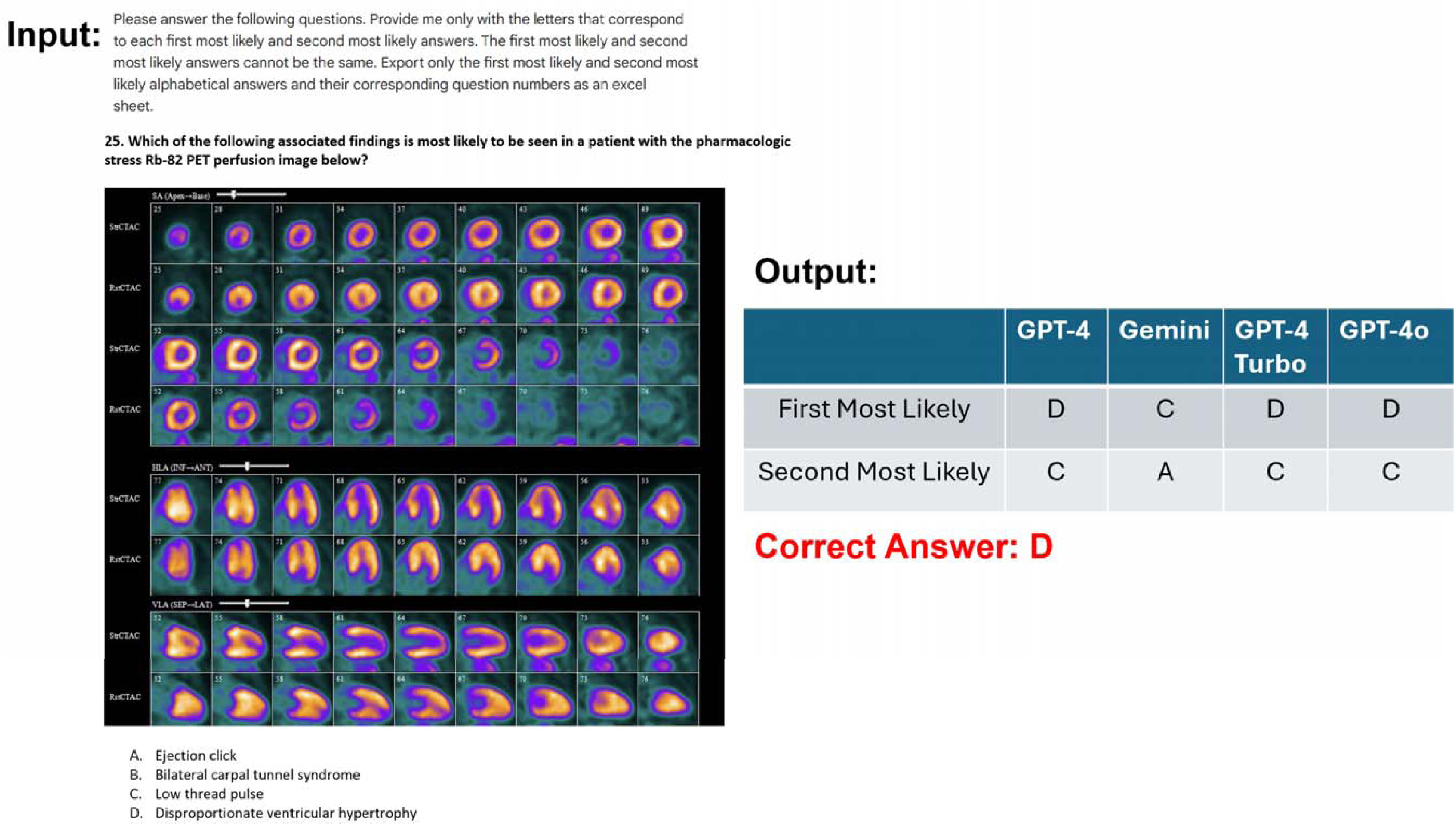
Section 3 Example Problem. The input and the three models’ output for a multimodal, text and image-based question from Section 3 (Test Selection, Stress and Nuclear Protocols Interpretation, Appropriate Use, and Risk Stratification) of the 2023 ASNC Board Preparation Exam. A first most likely and second most likely answer was provided by the models. Only the first most likely answer was scored and compared to the correct answer. GPT-4, GPT-4 Turbo and GPT-4o were able to answer correctly, whereas Gemini provided incorrect answers for 17 of 30 attempts. ASNC – American Society of Nuclear Cardiology

### Time progressive testing

Figure 4 shows overall results for time progressive manual testing of singular attempts taken 6 weeks apart in April 2024 and June 2024 of GPT-4, Gemini and GPT-4 Turbo. Although the models showed changes in total exam performance, none were significant: p=0.451 for GPT-4, p=0.377 for Gemini and p=0.089 GPT-4 Turbo. Sectional analysis revealed a significant degradation only in the performance of GPT-4 Turbo on Section 4, where GPT-4 Turbo answered 74.2% of answers correct in April 2024 and 54.5% of answers correct in June 2024 (p=0.023) (**Supplemental Table 1**). There were no significant differences in time progressive testing of image-based or text-only questions for the three models (**Supplemental Table 2**).

**Figure 4.**
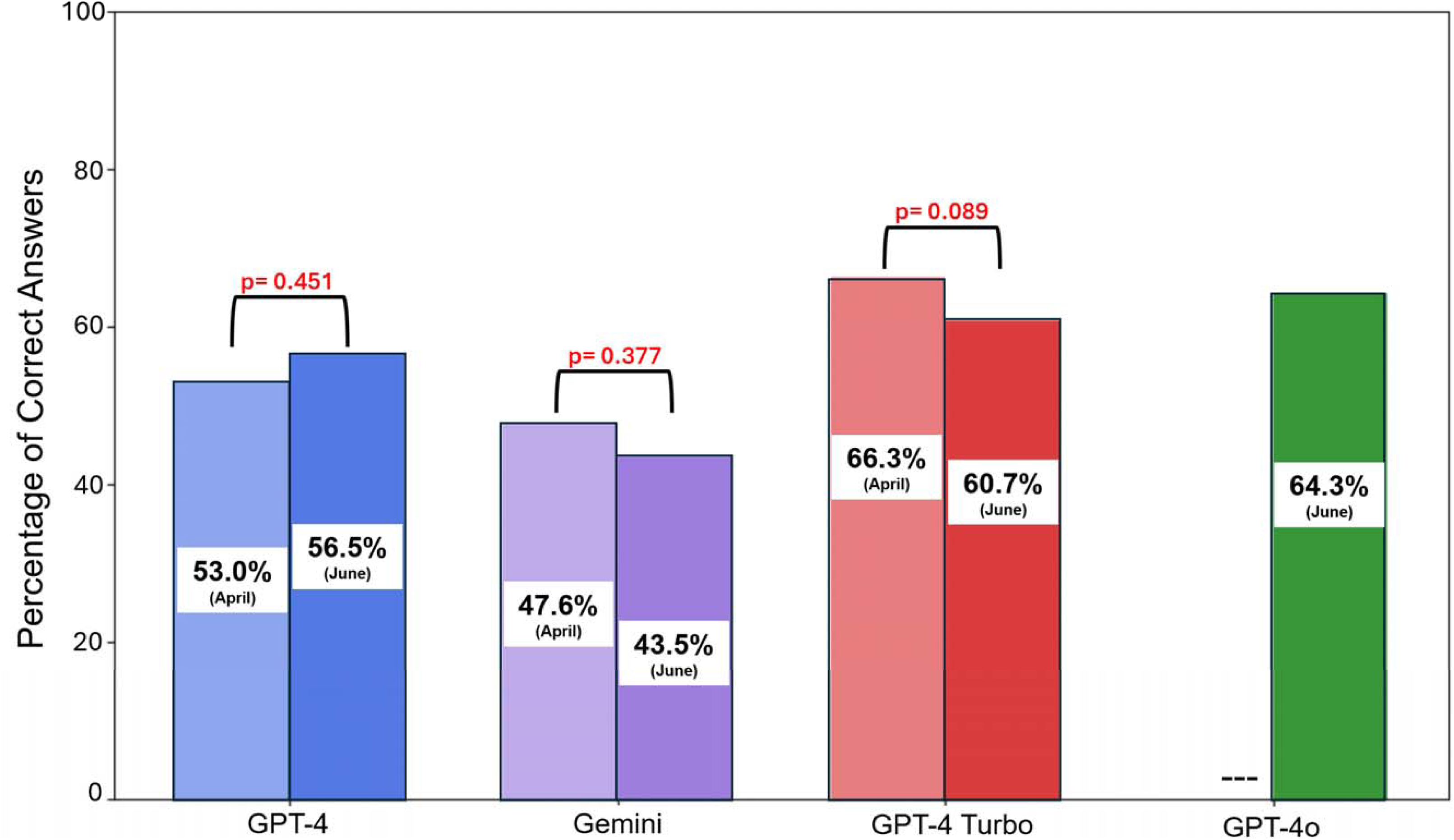
Time progressive analysis of overall performance of GPT-4, Gemini, and GPT-4 Turbo. The three models were tested on all 168 questions of the 2023 American Society of Nuclear Cardiology (ASNC) Board Preparation Exam six weeks apart in April 2024 and June 2024 for singular test attempts. GPT-4o was excluded from this analysis as it was released on May 13, 2024, after the first stage of data collection had commenced.

## DISCUSSION

LLMs are rapidly becoming used in all aspects of medicine by physicians and patients. We demonstrated for the first time the capability of these tools in handling a large number of advanced cardiovascular imaging subspecialty board questions, which include images. It is critically important for physicians to understand the capabilities and current limitations of such systems. The results of our comparative analysis among four leading LLMs – GPT-4, Google Gemini, GPT-4 Turbo and GPT-4o – on the 2023 ASNC Board Preparation Exam provide insights into their capabilities and limitations within the specialized field of nuclear cardiology and cardiac imaging in general.

The board preparation examination, designed to reflect the content outline of the CBNC exam, revealed a variance in performance across several analyzed categories. GPT-4o outperformed the other three models, achieving a significantly higher overall success rate. This superiority was most notable on text-only test questions and on Sections 1 and 4 of the exam, where GPT-4o’s performance significantly surpassed GPT-4, Gemini, and GPT-4 Turbo. However, GPT-4 Turbo also demonstrated strengths on certain portions of the exam, performing similarly to GPT-4o in Section 2 and outperforming all three other models in Section 3. Gemini was significantly weaker in answering image-based questions than the three other models, which all showed comparable results in this category. There were no significant differences in model performance over a 6-week time interval, with the exception of the degradation in GPT4-Turbo’s performance in Section 4. These outcomes suggest that, when considering overall capability and performance, GPT-4o’s algorithms may be better suited for handling the complexity and specificity of questions and tasks required of certified nuclear cardiologists.

This study is one of the first to include both multimodal, image- and text-based questions within a medical subspecialty domain, and the first in cardiovascular imaging to compare the performance of multiple LLMs in this capacity. GPT-3.5 and GPT-4 have only been tested on text-based cardiac imaging questions derived from common terminology used in radiology reports and cardiac imaging guidelines^24^. To our knowledge, previous studies have not yet examined the time-progressive performance of multiple multimodal compatible LLMs on medical subspecialty board exams, nor have they assessed GPT-4o. Previous research has explored GPT-4’s ability to answer hybrid image and text-based questions on the Japanese Emergency Medicine Board Exam^12^ and the American Shoulder and Elbow Surgeons Maintenance of Certification Exam^11^. GPT-4 with Vision (GPT-4V) was tested on the Japanese Otolaryngology Board Exam^10^, and GPT-4V Turbo was tested on the Japanese Diagnostic Radiology Board Exam ^14^. The multimodal capabilities of Bard (former Google Gemini) were previously evaluated on ophthalmology board exam practice questions^13^. Suh et al. investigated the ability of GPT-4V and Gemini Pro Vision to generate differential diagnoses for cases that included input images compared with expert radiologists, including twelve cases in the cardiovascular subspecialty^25^.

The scoring method used by the CBNC for the certification exam could not be precisely replicated in this study. The CBNC uses a criterion-referenced test, which bases scores on a set standard rather than comparing candidates’ performances against one another. Their “standard setting” process uses the Angoff Method, where a panel estimates the percentage of minimally competent candidates who would correctly answer each question^26^. The estimates are summed across questions for each judge, and the average across all judges determines the test’s cut-score^26^. The CBNC exam results are given as either a PASS or FAIL (with no set percentage required to pass). However, the CBNC does assign section evaluations as “Requires Further Study” for scores 60% or below and “May Require Further Study” for scores of 61-79%, suggesting that 60% is a meaningful threshold. Previous studies assessing the performance of LLMs on medical subspecialty board exams have reported passing thresholds ranging from 60%-70%^6–9,12,13^. Therefore, considering that correctly answering around 65% suggests a high likelihood of passing the CBNC exam, none of the models would have been highly likely to pass the overall exam. If we consider only text-based questions that do not include or require interpretation of images, GPT-4o would have been highly likely to pass.

Cardiovascular imaging and nuclear cardiology examinations reflect a particularly difficult challenge for LLMs given the combination of textual and image-based knowledge required. In this regard, we found that there was a decline in the accuracy of all four models for image-based questions compared to their performance for non-image-based questions. This observation aligns with previous findings by Yan et al. that reported that GPT-4V and Gemini Pro performed worse than random guessing on medical imaging diagnostic questions^27^. Additionally, our results revealed that Gemini performed significantly worse than the three GPT models on questions that include images, while the GPT models showed no difference in performance between their different versions (GPT, GPT-4 Turbo, GPT-4o). This suggests that Gemini 1.0 Pro with Vision is currently inferior in its medical image interpretation capabilities and that these capabilities have not improved between the developments of GPT, GPT-4 Turbo, and GPT-4o. Google, however, has recently introduced Med-Gemini, a new family of highly capable multimodal models specialized in medicine, which have demonstrated superior performance with medical image challenges compared to several LLMs^22,28^. Med-Gemini has not yet been released for general use and was not available for this study. Generally, to enhance the accuracy of LLMs on textual and image-based medical exam-style questions, further specialized in-context embedding retrieval method focused on medical images may be essential^29^, as models’ capabilities on exams appear to stem primarily from pre-training processes rather than methods such as exam-taking strategies or random guessing^16^. It is possible that only through such targeted improvements can LLMs hope to reach the interpretative proficiency of human experts, particularly in fields as specialized as nuclear cardiology, where the integration of multimodal data is crucial for accurate diagnostics.

Lastly, we observed that the models generally showed lower median percentiles in Section 2 (Acquisition and Quality Control, Gated SPECT, Artifact Recognition, and MUGA), where knowledge is typically sourced from specific nuclear cardiology literature. In contrast, median scores were generally higher in Section 1 (Radiation Safety), where information is available from a broader range of sources, and Section 3 (Test Selection and Appropriate Utilization), which is covered in general cardiology literature. Therefore, enhancing training sets, with highly specialized knowledge not readily available on the internet, will likely improve LLM performance in these areas where answers are derived from more obscure sources.

### Limitations

This study has several limitations. First, an official passing rate benchmark for this exam is not available, though it is estimated that between 60 and 65% of correct answers would translate into a high likelihood of passing the CNBC, as discussed earlier. Additionally, we utilized the ASNC board preparation examination as a surrogate for the CBNC exam. While our results might not directly correlate with performance on the CBNC exam, the ASNC board preparation examination is frequently used by physicians to gauge their readiness. GPT-4o was not included in time-progressive performance testing since it was released on May 13, 2024, subsequent to the commencement of this study phase in April 2024. Future studies could explore the time-progressive performance of GPT-4o. Lastly, due to ongoing enhancements in LLMs, the responses from the three chatbots could evolve over time, possibly leading to different answers if the same queries were posed again in the future.

## CONCLUSION

GPT-4o demonstrated superior performance compared to three other LLMs and achieved an overall score that is most likely within or just outside the range required to pass a test akin to the CBNC examination. Although improvements are still needed in its ability to accurately interpret medical images, these results suggest GPT-4o’s potential for supporting physicians in answering a wide range of questions related to nuclear cardiology field.

## Data Availability

All data produced in the present study are available upon reasonable request to the authors

## Abbreviations

GPT: Generative Pre-trained Transformer
LLM: Large Language Model
CBNC: Certification Board of Nuclear Cardiology
ASNC: American Society of Nuclear Cardiology
SPECT: Single Photon Emission Computed Tomography
PET: Positron Emission Tomography

---

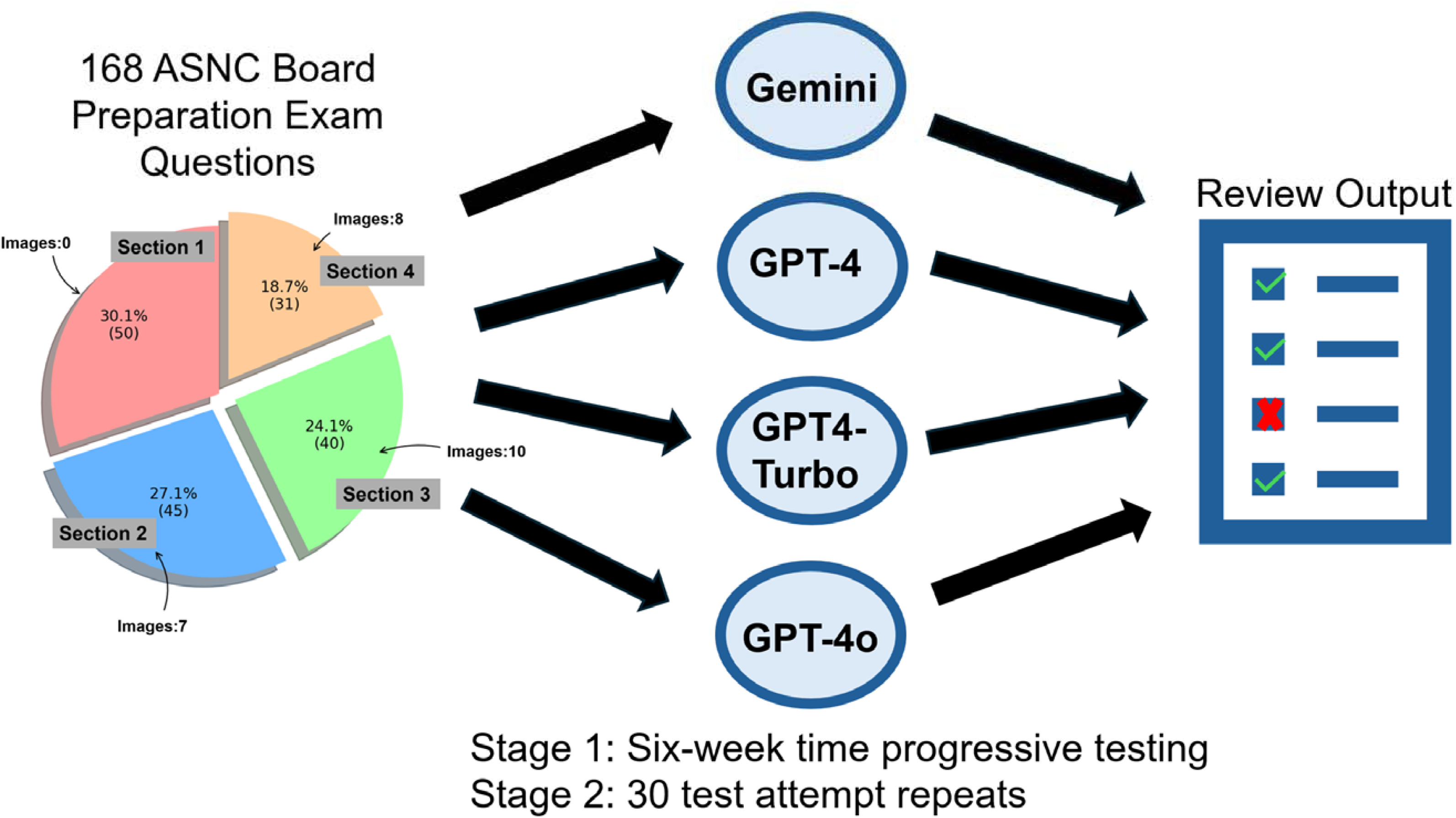
Central Illustration. Study Overview. The pie chart shows the breakdown of the percentage and (total count) of questions in each section of the exam. ASNC – American Society of Nuclear Cardiology

