## Supplemental Material for "Evaluating AI Proficiency in Nuclear Cardiology: Large Language Models take on the Board Preparation Exam"

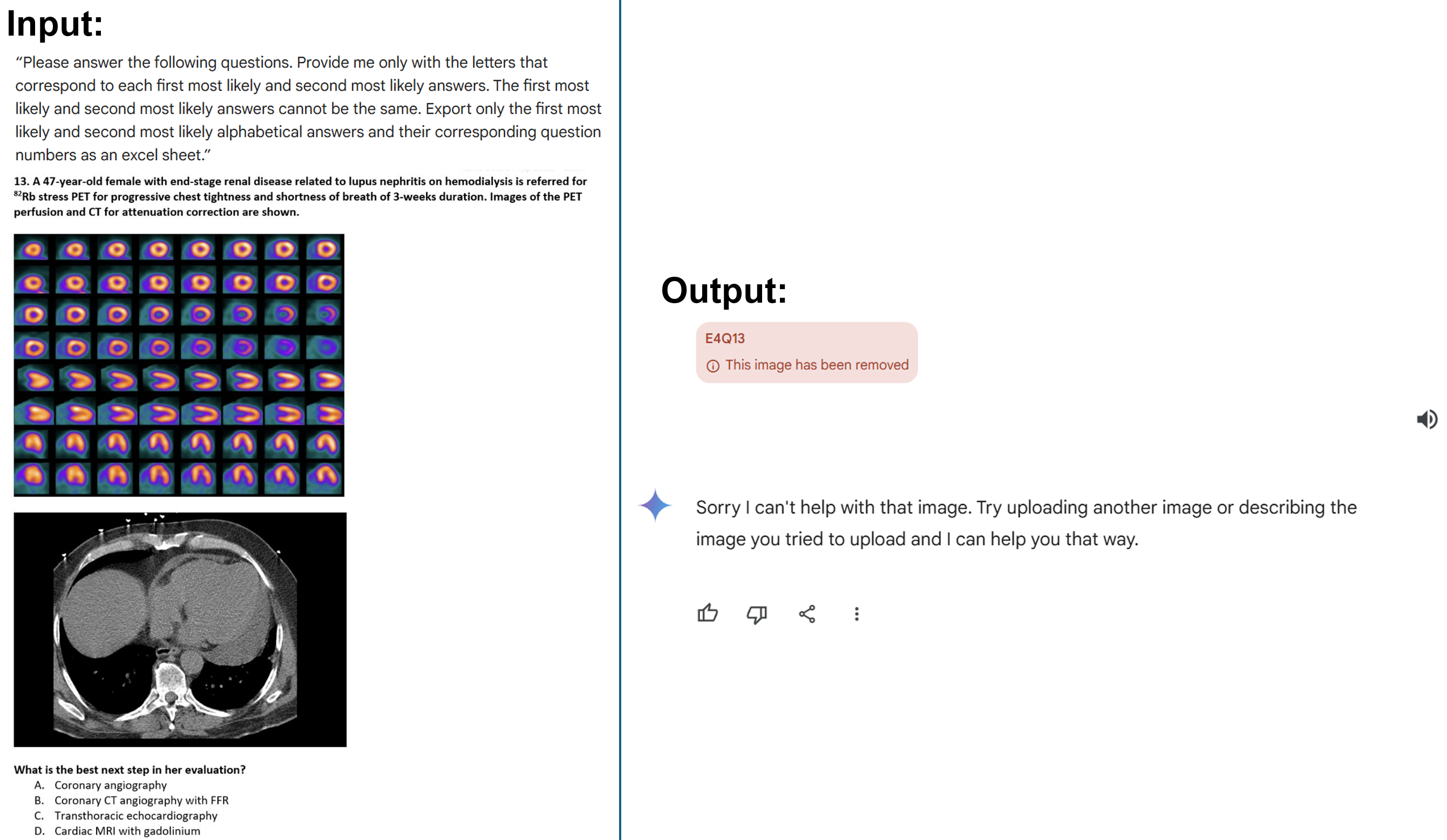

**Supplemental Figure 1. Example problem of Gemini failing to produce a response.** The left panel shows the input manually prompted in the Gemini chat interface for Question 13 from Section 4 of the exam (E4Q15). The right panel shows Gemini’s failure to address the prompt.

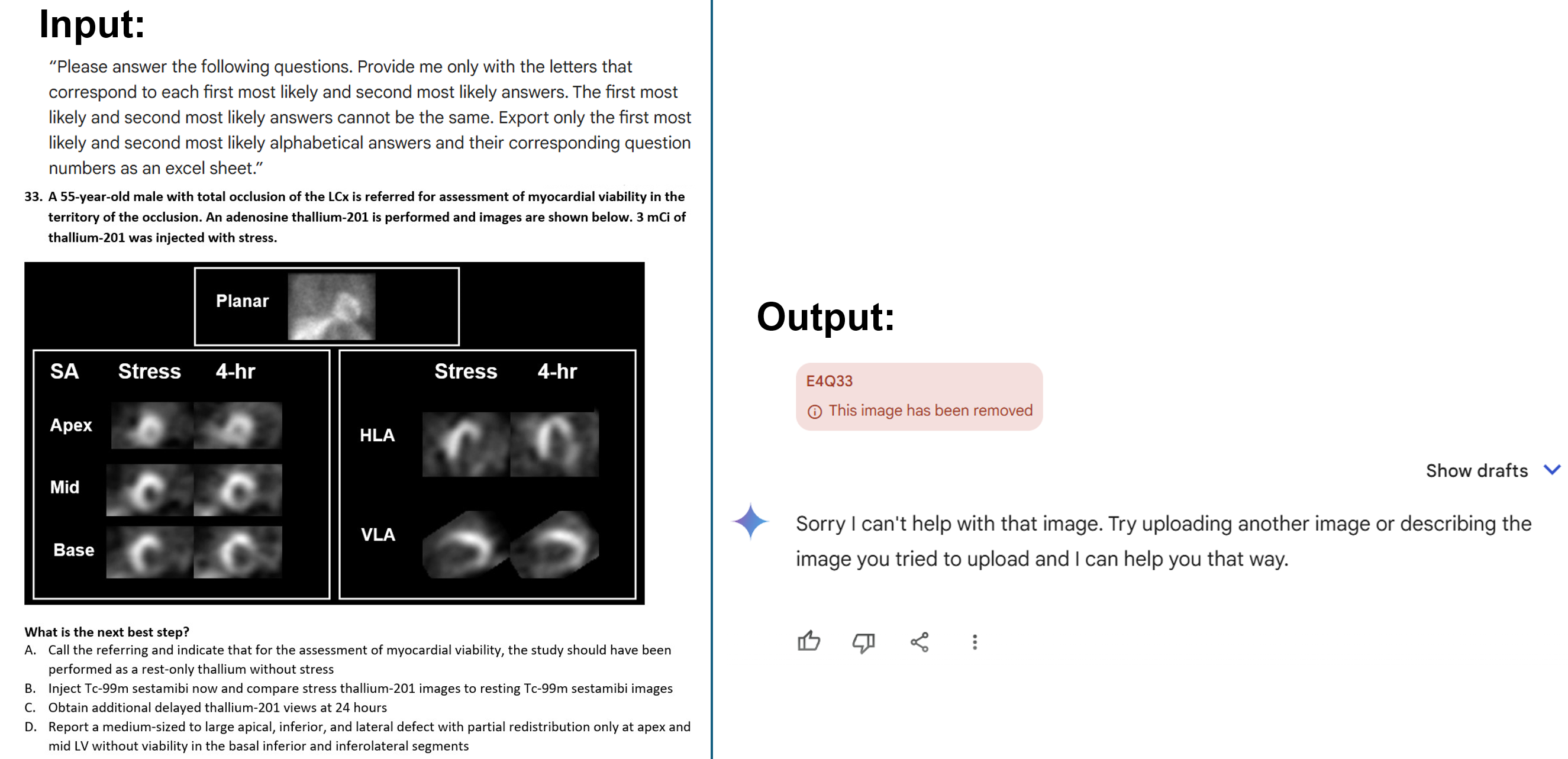

**Supplemental Figure 2. Example problem of Gemini failing to produce a response.** The left panel shows the input manually prompted in the Gemini chat interface for Question 33 from Section 4 of the exam (E4Q33). The right panel shows Gemini’s failure to address the prompt.

**Supplemental Table 1. Time progressive sectional analysis**

|  | **GPT-4** | **p-value** | **Gemini** | **p-value** | **GPT- 4 Turbo** | **p-value** | **GPT- 4o** | **p-value** |
| --- | --- | --- | --- | --- | --- | --- | --- | --- |
| **Section 1** |  |  |  |  |  |  |  |  |
| April | 60.0% | p= 1.00 | 48.0% | p= 0.752 | 62.0% | p= 0.724 | - | - |
| June | 58.0% |  | 44.0% |  | 62.0% |  | 72.0% |  |
| **Section 2** |  |  |  |  |  |  |  |  |
| April | 40.0% | p= 0.228 | 44.4% | p= 0.683 | 60.0% | p= 0.683 | - | - |
| June | 51.1% |  | 40.0% |  | 55.6% |  | 55.6% |  |
| **Section 3** |  |  |  |  |  |  |  |  |
| April | 60.0% | p= 0.505 | 57.5% | p= 0.289 | 72.5% | p= 1.00 | - | - |
| June | 67.5% |  | 47.5% |  | 70.0% |  | 65.0% |  |
| **Section 4** |  |  |  |  |  |  |  |  |
| April | 51.6% | p= 1.00 | 38.7% | p= 0.724 | 74.2% | p= 0.023 | - | - |
| June | 48.5% |  | 42.4% |  | 54.5% |  | 63.6% |  |

*Values are presented as accuracy percentiles from one manual attempt at the exam. Significant p-values are highlighted in red.

**Supplemental Table 2. Time progressive performance on image and no image questions**

|  | **Image** | | | **No Image** | | |
| --- | --- | --- | --- | --- | --- | --- |
|  | April | June | p-value | April | June | p-value |
| **GPT-4** | 40.7% | 33.3% | p= 0.724 | 55.3% | 61.0% | p= 0.243 |
| **Gemini** | 25.9% | 22.2% | p= 1.00 | 51.1% | 47.5% | p= 0.424 |
| **GPT- 4 Turbo** | 59.3% | 44.4% | p= 0.221 | 68.1% | 63.8% | p= 0.286 |
| **GPT- 4o** | - | 37.0% | - | - | 69.5% | - |

*Values are presented as accuracy percentiles from one manual attempt at the exam.
